# Direct-acting antivirals for chronic hepatitis C infection: a protocol for a systematic review of observational studies

**DOI:** 10.1101/2024.10.21.24315594

**Authors:** Buddheera W.M.B. Kumburegama, Andreas T. Kristensen, Goran Bjelakovic, Dimitrinka Nikolova, Mark A. Asante, Milica Bjelakovic, Ronald L. Koretz, Mithuna M. Balakumar, Martin E. Michelsen, Sarah L. Klingenberg, Christian Gluud

## Abstract

**Background:** Chronic hepatitis C virus infection presents a substantial global health burden, frequently resulting in severe liver conditions. Hepatitis C virus (HCV) therapy requires complex decision-making. Direct-acting antivirals (DAAs) offer a potential solution by targeting viral proteins to inhibit replication. Understanding DAAs real-world effectiveness and how they impact long-term outcomes beyond clinical trials is essential. We aim to comprehensively evaluate the benefits and harms of DAAs in individuals with chronic HCV infection, reported in observational studies.

**Methods:** We will consider for inclusion prospective and retrospective observational studies with quasi-randomised, cohort, case-control, controlled before-and-after, and cross-sectional designs. Our experimental interventions will be any class of DAAs available on the market or in development. DAAs could have been administered alone, in combination, or with other medical co-interventions. Our control interventions will be untreated chronic HCV conditions, with or without placebo.

Participants will be adults, regardless of demographics, treatment history, or healthcare setting. Our primary outcomes will be participants experiencing hepatitis C-related morbidity or all-cause mortality, serious adverse events, and health-related quality of life. Secondary outcomes will include all-cause mortality, cirrhosis, variceal bleeding, hepato-renal syndrome, hepatocellular carcinoma, hepatic encephalopathy, non-serious adverse events, liver transplantation, lack of sustained virological response, histological improvement, and decreases in alanine aminotransferase and aspartate aminotransferase levels.

We will apply search strategies to search MEDLINE, Embase, Web of Science, grey literature, and trial registers. We will use Covidence^®^ to screen the result, including citations. Individual double-data extraction will include study details and outcomes, with independent review authors resolving discrepancies. We will assess bias using the ROBINS-I tool. Meta-analyses will employ random-effects models for both dichotomous and continuous outcomes, assessing heterogeneity. Subgroup and sensitivity analyses will explore effect modifications and address missing data. Trial Sequential Analysis will control type I and type II errors. We will evaluate publication bias using funnel plots and Egger’s regression test and assess certainty of evidence using GRADE.

**Discussion:** The findings will inform clinical decisions and benefit those affected by HCV, healthcare professionals, and policymakers.

**Systematic review registration:** PROSPERO: CRD42023494844

## Background

### Description of the condition

Hepatitis C virus (HCV) was identified as the agent responsible for liver infection in most patients with ‘non-A, non-B hepatitis’ as late as 1989 [38]. Since then, HCV has been recognised as a major cause of liver cirrhosis and hepatocellular carcinoma [13,25]. Globally, an estimated 58 million people are infected with chronic HCV, with an additional 1.5 million people acquiring the infection each year [47,83]. In 2019, HCV was responsible for approximately 290.000 deaths, mostly due to cirrhosis or hepatocellular carcinoma [16,19,83]. In 2020, the global prevalence of HCV infection was estimated at 0.7% (95% uncertainty interval 0.7% to 0.9%) [19,61].

HCV is an enveloped single-stranded positive-sense ribonucleic acid (RNA) virus [21,69]. The genome of the HCV contains an open reading frame encoding a polyprotein [21,69]. This polyprotein is processed to yield structural (core, glycoproteins E1, and E2, and protein P7) and non-structural (NS2, NS3, NS4A, NS4B, NS5A, and NS5B) proteins [13,21,69].

Classification of HCV is based on phylogeny (i.e. history of evolution) and sequence diversity, which defines eight major HCV genotypes and around 90 subtypes [9,34]. The geographical distribution and the prevalence of the eight genotypes varies [9,49].Genotype 1 is highly prevalent, accounting for 46% of infections around the world[49,69]; genotype 2 dominates in West Africa; genotype 3 in South Asia and parts of Scandinavia; genotype 4 in Central and North Africa; genotype 5 in South Africa; and genotype 6 and 7 in South-East Asia [30,49,69]. Genotype 8 is genetically distinct from genotype 1to7 and has been detected in four people who were originally from Punjab, India, but now reside in Canada and are not epidemiologically linked [9]. HCV is primarily transmitted through parenteral exposure to contaminated blood, such as needle sharing among people who inject drugs. However, HCV can also be transmitted by sharing personal care items that have been in contact with blood, vertical transmission from mother to child during childbirth, unprotected sexual contact, occupational exposure to blood, and others [27,73].

The signs and symptoms of HCV are similar across genotypes, though genotype 3 is associated with increased risks of hepatic steatosis and progressive liver disease [69]. Notably, a large percentage of individuals with chronic HCV infection may not progress to severe liver complications. HCV infection is often asymptomatic, and in individuals where the disease does not progress to cirrhosis or hepatocellular carcinoma, infection might not cause immediate harm [44]. Approximately 20% of people infected with HCV have self-limited acute hepatitis [44], but in the remaining 80%, the virus leads to a chronic infection [83]. A systematic review of 111 studies analysing the natural history of HCV infection found that the prevalence of cirrhosis 20 years after HCV infection was 16% [77]. Other studies have reported that progression into cirrhosis occurred in approximately 20% [15,44,80]. Approximately 10%-20% of people with chronic HCV infection progress to end-stage liver disease (i.e. decompensated cirrhosis or hepatocellular carcinoma), which corresponds to 8%-16% of people infected with HCV [44,80].

The host’s genetic factors, such as the interleukin-28 beta (IL-28B) subunit gene, is associated with increased risk of HCV outcomes, affecting both the potential for spontaneous viral clearance and the response to interventions with pegylated interferon α (peg-IFNα) and ribavirin [6].IL-28B positivity is associated with higher rate of spontaneous viral clearance and improved viral clearance following intervention with peg-IFNα and ribavirin treatment [6,41,51].

Treatment with peg-IFNα and ribavirin compared with other non-direct acting antiviral drugs or no intervention increased the proportion of people with sustained virologic response (SVR) defined as aviraemia12 or 24 weeks after the end of antiviral treatment [25,60]. Treatment with peg-IFNα and ribavirin is associated with serious adverse events and often leads to treatment discontinuation [10,33,44,45,65]. Furthermore, the impact of treatment on patient-centred outcomes (i.e. quality of life, HCV-related complications, or mortality) remain unclear [10,33,35,44,45,65]. While treatment with peg-IFNα and ribavirin improve the rate of SVR compared with or to other non-direct-acting antiviral drugs, it is important to note that findings from the HALT-C trial and the study by Koretz and colleagues have raised questions about potential adverse events and an association with increased mortality despite a higher rate of SVR [20,45].The risks of adverse events associated with IFN and ribavirin treatment spurred the development of alternative interventions, such as the direct-acting antivirals (DAAs).

### Description of the intervention

DAAs are compounds designed to selectively act on the non-structural (NS) viral proteins, leading to the interruption of viral replication to inhibit infection [60,63].There are four classes of DAAs, each classified by the mechanism of action and therapeutic target: non-structural 3/4A (NS3/4A) protease inhibitors (PIs); NS5B nucleoside polymerase inhibitors (NPIs); NS5B non-nucleoside polymerase inhibitors (NNPIs); and NS5A protein inhibitors [47,60,63]. Table 1 provides a summary of various DAAs and their current market status [25].

**Table 1.** Types of DAAs against hepatitis C virus and their relation to the market. The table presents a list of 61 DAAs, categorized by their class. If a DAA lacks a generic or brand name, we refer to it by its experimental compound number prefix. NS3/NS4A inhibitors: Non-structural protein 3/4A protease inhibitors NS5B inhibitors: Non-structural protein 5B polymerase inhibitors NPI: Nucleotide polymerase inhibitors NNPI: Non-nucleotide polymerase inhibitors NS5A inhibitors: Non-structural protein 5A inhibitors

| DAAs currently in the market |  |  |  |
| --- | --- | --- | --- |
| NS3/NS4A inhibitors | NS5B inhibitors |  | NS5A inhibitors |
|  | NPI | NNPI |  |
| Asunaprevir | Sofosbuvir | Dasabuvir | Daclatasvir |

**Table 1. Types of DAAs against hepatitis C virus and their relation to the market.**
| <u>Glecaprevir</u> |  |  | Elbasvir |
| --- | --- | --- | --- |
| Grazoprevir |  |  | Ledipasvir |
| Paritaprevir |  |  | Ombitasvir |
| Voxilaprevir |  |  | Pibrentasvir |
| Danoprevir |  |  | Velpatasvir |
|  |  |  | Ravidasvir |
| **DAAs under development** |  |  |  |
| NS3/NS4A inhibitors | NS5B inhibitors |  | NS5A inhibitors |
|  | NPI | NNPI |  |
| ACH-2684/Deldeprevir/Neceprevir | ALS2200/VX135 | ABT-072 | ACH-2928 |
| Celuprevir | BILB1941 | Beclabuvir | GSK2336805 |
| Faldaprevir | GS0938/PSI352938 | BI201127 | MK-8408/<br>Ruzasvir |
| GS9256 | GS6620 | Deleobuvir | Odalasvir |
| IDX320 | GS9851(PSI 7851) | Filibuvir | PPI461 |
| Narlaprevir | IDX184 | GSK2878175/GSK175 | Ravidasvir |
| PHX1766 | INX189/BMS986094 | IDX375 | Samatasvir |
| Sovaprevir | Mericitabine | MK-3281 |  |
| Vaniprevir | MK3682<br>/uprifosbuvir | Nesbuvir |  |
| Vedroprevir |  | Radalbuvir |  |
|  |  | Setrobuvir |  |
|  |  | Tegobuvir |  |
|  |  | TMC-647055 |  |
|  |  | VCH-759 |  |
|  |  | VCH-916 |  |
|  |  | VX222 |  |
| **DAAs withdrawn from the market** |  |  |  |
| NS3/NS4A inhibitors | NS5B inhibitors |  | NS5A inhibitors |
|  | NPI | NNPI |  |
| Boceprevir |  |  |  |
| Simeprevir |  |  |  |
| Telaprevir |  |  |  |

### NS3/4A protease inhibitors

The NS3/4A protease inhibitors telaprevir and boceprevir, were approved for chronic genotype 1 HCV infection in 2011[26,54]. Treatment with a protease inhibitor combined with peg-IFNα and ribavirin resulted in SVR of 68% to 75% in treatment-naive patients and 59% to 88% in treatment-experienced patients [69,65].Drawbacks associated with the use of telaprevir or boceprevir encompassed a rapid development of viral resistance [15], long treatment duration (24-48 weeks), and an increased risk of serious adverse events [15,65,69].For these reasons and with the introduction of second-generation protease inhibitors, telaprevir was withdrawn from the market [40,65], and boceprevir ceased to be recommended as a treatment option [65].

Simeprevir and paritaprevirare distinguished by their theoretically high potency, susceptibility to the development of viral resistance, and cross-resistance (drug-drug interaction) with other NS3/4A protease inhibitors. Regrettably, the presence of cross-resistance can lead to a decrease in the effectiveness of these drugs [66].

In 2013, regulatory approval was granted for the use of simeprevir in combination with peg-IFNα and ribavirin [24]. Simeprevir has demonstrated efficacy against HCV genotypes 1, 2, 5, and 6 and is generally associated with manageable adverse effects [15,24]. The typical treatment duration for simeprevir is 24 weeks. Paritaprevir is often co-administered with low-dose ritonavir, an antiretroviral protease inhibitor commonly used in HIV/AIDS treatment, to achieve a pharmacologic boosting effect [60]. Paritaprevir and ritonavir are also offered in combination with ombitasvir (an NS5A inhibitor) and are commonly prescribed with the NNPI dasabuvir (see below) [60].

### NS5Bpolymerase inhibitors and NS5A inhibitors

NS5B polymerase inhibitors have been used across various HCV genotypes and are characterised by their substantial theoretical potency and a high resistance barrier, owing to the highly conserved active site in NS5B protein across different HCV genotypes [15,24,65].

The NS5B polymerase inhibitors can be classified into NPIs and NNPIs. Sofosbuvir, the first NPI approved in 2013, is well-tolerated [65,66]. Sofosbuvir is typically administered once daily for 12 weeks, often as part of a combination therapy for HCV. Notably, it exhibits a limited cross-resistance interaction profile when compared to earlier DAAs [65,66]. NNPIs, such as dasabuvir, interact with regions on the NS5B polymerase that are less crucial for viral survival. As a result, dasabuvir has the lowest theoretical barrier to resistance among NS5B polymerase inhibitors [66].

Due to the theoretically limited resistance potential, NS5A inhibitors are recommended alongside combination agents, including NS3/4A inhibitors, and NS5B polymerase inhibitors [15]. The NS5A inhibitors daclatasvir, ledipasvir, and ombitasvir were approved for use in the European Union in 2014 and in USA in 2015, for use in people with chronic HCV infection in combination with other DAAs [65,79].

### How the intervention might work

DAAs are compounds designed specifically to target non-structural proteins encoded by HCV to disrupt viral replication and progression of the infection [8,60]. The efficacy of DAAs may vary depending on the HCV genotype and subtype [8,60].

### Why it is important to do this review

Previous randomised clinical trials examining the impact of DAAs have predominantly centred on evaluating SVR as the primary outcome. SVR is characterised by the absence of detectable virus in the blood either 12 or 24 weeks after the end of antiviral therapy [40,57,62]. The SVR rates varied with the specific DAA regimens used in the trials, irrespective of the treatment duration, previous treatment status, or viral genotype. In general, these rates were high (range 67%-99%) [43]. Therefore, the overall results suggest that DAAs are more effective in achieving SVR than peg-IFNα and ribavirin [1,4,40,46,48,62,84]. We obtained a similar result in our systematic review of 138 randomised clinical trials including 25,232 participants on DAAs versus placebo or no intervention [41]. The trials included in the review had short-term follow-up, designed primarily to assess the effect of treatment on SVR with an average duration of 34 weeks. We found that DAAs might reduce the risk of no SVR to 23.8% compared with 54.1% in participants treated with placebo or no intervention (relative risk [RR] of no SVR0.44, 95% confidence interval [CI]0.37-0.52; 6,886 participants; 32 trials; low certainty evidence) [41]. However, the meta-analyses of DAAs on the market or under development versus placebo or no intervention did not show evidence of a difference in either mortality (DAA 15 events /2377analysed[0.63%] versus control 1/617 [0.16%]; 11 trials; odds ratio [OR] 3.72, 95% CI 0.53 - 26.18;low certainty evidence) or serious adverse events (DAA 373/7251[5.2%] versus control125/2243[5.6%]; 43 trials; OR 0.93, 95% CI 0.75 −1.15;very low certainty). Furthermore, we found no data on any hepatitis C-related morbidity [41]. Since 2017, new trials exploring DAAs for chronic hepatitis C have emerged. Recognising these developments, we are presently updating our systematic review [41].

While SVR is often seen as a valid patient-centred outcome associated with improved survival, it is essential to subject this association to careful analysis [29]. Supporting evidence primarily stems from studies comparing treated patients who achieved SVR to those who did not (i.e. observational data). Since all patients in these studies received the same treatment, it is crucial to recognise that any differences in outcomes between the SVR and non-SVR groups cannot be attributed to the treatment alone. Therefore, it is essential to employ caution when using such studies as supporting evidence. Acknowledging the correlation between SVR and improved survival, it is vital to assess the treatment efficacy in achieving relevant outcomes and to consider patient-specific factors [29].

Observational studies have reported associations between SVR, increased survival, and fewer liver-related complications [14,55]. This may be attributed to eradication of the HCV and stabilisation or reversal of fibrosis [25]. However, such associations cannot establish causation [11,29,52]. As described above, establishing a direct relationship between SVR and a favorable clinical outcome has not yet been confirmed through randomised clinical trials, in which causation may be established 11,29,41,52]. The clinical effects of DAAs, therefore, remain unclear and questioned [41,44]. It is crucial to note that SVRs are not randomly achieved but are influenced by various prognostic factors. These factors include infection duration, severity of liver fibrosis, HCV-RNA levels, presence of the IL28B gene, sex, race, and body mass index [3,56]. While randomised clinical trials are often regarded as the gold standard for establishing causation, observational studies play a crucial role in assessing real-world effectiveness and long-term outcomes in diverse patient populations. Furthermore, it is important to highlight that no systematic review of observational studies, considering the risks of systematic, design, or random errors has been published in this setting [36,42].

Observational studies have the advantage of potentially offering long-term follow-up on outcomes that may not be feasible to explore in a randomised clinical trial. All observational studies and their various designs, however, have serious limitations, among which are confounding, lack of a control group, limited generalisability, incomplete data, high risks of publication bias, and other biases [28,64,76] Ultimately establishing causality can be difficult [12,14].

### Objective

To assess the benefits and harms of DAAs in people with chronic HCV infection as assessed in observational studies.

## Methods

### Criteria for considering studies for this review Types of studies

We will include observational studies that evaluated the benefits and harms of DAAs in participants with chronic HCV infection. We will include the following observational designs:

- quasi-randomised studies
- cohort studies
- case-control studies
- controlled before-and-after studies
- cross sectional studies

Both prospective and retrospective designs will be eligible. There will be no minimum duration requirement for the studies regarding treatment or follow up.

### Types of interventions

Any DAA drugs (described in Table 1) currently on the market or on the way to the market.

### Experimental intervention

The experimental interventions include the administration of any DAA drug (described in Table 1) either as a single agent, in combination with another DAA, or combined with other medical co-interventions.

### Control intervention

The control interventions are defined as untreated chronic HCV conditions with or without placebo.

### Types of participants

We will include studies involving adults (as defined by the researchers) of any sex, ethnicity, occupation, or residency, who have been diagnosed with chronic HCV infection. These studies may involve treatment-naive or treatment experienced participants.

We will not exclude participants based on any HCV-related comorbidity, including concurrent HIV infection, hepatitis B infection, alcoholism, other specific comorbid conditions. We will allow pregnant women and individuals with a history of drug use, including the use of injection drugs.

We will include studies conducted in any healthcare setting, including hospitals, clinics, primary care settings, and country.

### Outcomes

#### Primary outcomes

- Proportion of participants with hepatitis C-related morbidity or all-cause mortality. Hepatitis C-related morbidity is defined as either cirrhosis, ascites, variceal bleeding, hepato-renal syndrome, hepatic encephalopathy, or hepatocellular carcinoma.
- Proportion of participants with one or more serious adverse events. We will use the International Conference on Harmonisation Guidelines for Good Clinical Practice’s (ICH-GCP) [39] definition for serious adverse events, that is any untoward medical occurrence that resulted in death, was life-threatening, required hospitalisation or prolongation of existing hospitalisation, or resulted in persistent or significant disability or incapacity [39]. We will also consider the definitions used by the study authors.
- Health-related quality of life (any validated continuous outcome scale used by the authors).

#### Secondary outcomes

- All-cause mortality.
- Proportion of participants with cirrhosis.
- Proportion of participants with ascites.
- Proportion of participants with variceal bleeding.
- Proportion of participants with hepato-renal syndrome*.
- Proportion of participants with hepatocellular carcinoma.
- Proportion of participants with hepatic encephalopathy*.
- Proportion of participants with adverse events considered non-serious (any adverse event not defined as a serious adverse event). We also plan to assess each non-serious adverse event separately.
- Proportion of participants who received liver transplantation.
- Proportion of participants without SVR: this outcome assesses the presence of detectable HCV RNA (above an author-defined lower limit of detection) in the serum by a sensitive polymerase chain reaction (PCR)-based assay or transcription-mediated amplification testing, during the 12-24 weeks following the completion of treatment.
- Proportion of participants without histological improvement*.
- Proportion of participants without significant reductions in alanine aminotransferase (ALT) and aspartate aminotransferase (AST) serum levels*.

### Data search

We will search MEDLINE, Embase, and Web of Science. Additionally, we will search the grey literature, including conference proceedings, theses, and dissertations, using online search engines like Scopus. To ensure the thoroughness of the review, we will use a multi-faceted approach that includes citation tracking and input from experts in the field. We will also search the reference lists of included studies, as well as trial registers. The search will be designed to capture both published and unpublished studies with no language restrictions.

We will use Covidence to remove duplicate records during our research process. We will then screen the results for relevance and ultimately include a final number of studies that meet the inclusion criteria.

Our review authors (WMBBK, ATK, MAA, MMB, MEM) will individually and collaboratively evaluate all retrieved records. If one author deems a study relevant while another does not, the authors will engage in a discussion. In the event that a consensus is not reached, CG will facilitate a resolution.

### Excluded studies

The screening process will be carefully documented, and the decisions made during this stage will be outlined in the final publication. Additionally, it will also include a link to the list of excluded studies which will be stored at Zenodo.

### Data collection

The data collection and extraction process will adhere to standard methodological procedures in accordance with Cochrane standards [36]. We will gather data on study design, setting, sample size, participant demographics (such as HIV infection status, chronic kidney disease, cryoglobulinemia, viral genotypes, genotypes of the IL28B gene(IL28B[CC], IL28B [CT], IL28B [TT], IL28B [CT + TT])[2,23], geographical region, and ethnicity), type of DAA, previous treatments status (including ribavirin and ribavirin), and median antiviral medication dose. We will also record data on treatment duration, follow-up, funding, and outcome reporting.

Our team of review authors will work independently and in pairs using an Excel sheet designed for this purpose. Any discrepancies that may arise during data extraction will be resolved through discussion and, if necessary, arbitrated by CG. In instances where relevant data is not available, direct contact with authors will be made for clarification.

### Risk of bias assessment

We will use the risk of bias in non-randomised studies of Interventions (ROBINS-I tool) [74] to assess potential biases in the included studies. Our focus is on understanding the impact of assignment to the intervention and will adhere to the intention-to-treat principle. ROBINS-I comprises seven domains, including participant selection, intervention classification, deviations from intended interventions, missing data, outcome measurement, selection of reported results, and overall risk of bias, assessing study design and implementation for potential biases.

For each domain, we will assess risk as low, moderate, serious, critical, or no information. Our review authors will work in pairs independently to assess risk of bias on an individual outcome basis. Our assessment will be based on the collective impression of the study rather than a predetermined algorithm, acknowledging variations across studies.

### Data synthesis

The studies included in this systematic review will be synthesised using a meta-analytic approach. The dichotomous and continuous outcomes in this review will be analysed as relative risks (RR) and mean differences (MD), respectively. These measures will be used to calculate the summary estimates of the effects of DAAs.

We will consider the bias risks of the studies in our analysis and perform a sensitivity analysis to investigate the effects of studies at the lowest risks of bias. We will assess outcomes at the ‘longest follow-up’.

For the health-related quality of life outcome, we will compare the different scales employed by the studies and determine if a standardised effect size (SMD) can be used to compare the results, along with calculating the associated 95% confidence interval (CI). If a standardised effect size is not applicable, we will report the results separately based on the specific measurement method used.

Subsequently, we will indicate the direction of ‘better’ and ‘worse’ for the results obtained with the different scales or the standardised effect size result. Specifically, for each outcome, we will provide interpretation guidance to clarify whether an increase or decrease in the reported values indicates an improvement or deterioration in health-related quality of life We will use a random-effects model to combine the estimates across studies, given the expected heterogeneity between studies. We will present the results in a forest plot.

### Heterogeneity assessment

In our protocol, we will assess heterogeneity across three dimensions: clinical, methodological, and statistical. In our clinical heterogeneity assessment, we will use the recently developed Clinical Diversity in Meta analyses (CDIM) tool, designed for the evaluation of clinical diversity in meta-analyses of interventions [5]. CDIM is a validated instrument comprising four domains, including settings, population characteristics, interventions, and outcomes. Our approach involves a two-step process where the first two authors independently assess clinical diversity within these domains. Subsequently, consensus scores will be achieved. The scoring system ranges from 0 to 22, with 0 indicating no clinical heterogeneity and 22 representing high levels of heterogeneity. Our planned subgroup analyses will explore potential effect modification by identifying risk factors from large observational studies.

In our methodological heterogeneity assessment, we will use the ROBINS-I tool to assess the risk of bias in studies. We will also use the GRADE approach to assess the quality of evidence of outcomes. We will use meta-regression to explore its potential sources and impact on our findings [18,68].

In our statistical heterogeneity assessment, we will use multiple methods, including visual inspection of forest plots and statistical tests. Visual inspection of forest plots is a recommended approach to identify substantial statistical heterogeneity [18,42,68]. We will also use the Chi-squared test to assess the presence of heterogeneity, with a significance level set at P value < 0.10 [17]. Additionally, we will measure the extent of heterogeneity using the I^2^ statistic [17,37,68].

### Assessment of publication bias

The selective publication of studies with positive results, also known as publication bias, can threaten the validity of a systematic review. To assess the potential impact of publication bias on the validity of results, we will use two methods: the funnel plot and Egger’s regression test [18,42,68]. However, it is important to note that funnel plots tend to be more reliable when there are enough studies, usually 10 or more [36]. With a smaller number of studies, the natural variation in results due to sample size can lead to an asymmetric funnel plot, making it challenging to distinguish actual publication bias and random variation [58]. Furthermore, for dichotomous outcomes, we will use the Harbord test to detect asymmetry when the between-study variance (τ^2^) is less than 0.1, and the Rücker test when τ^2^ is equal or greater than 0.1 [36,58,67]. For the continuous outcome (health-related quality of life), we will use the regression asymmetry test [22]. This approach will enable us to identify associations rather than causal relationships regarding the impact of DAAs on SVR and clinical outcomes among individuals with HCV infection. In case we detect any signs of publication bias, our discussion will encompass the potential implications of such bias on our findings and conclusions.

### Subgroup analysis

Based on the characteristics of the participants and studies, we will conduct subgroup analyses to explore potential associations regarding the use of DAAs with SVR and clinical outcomes in individuals with HCV. To ensure the reliability of our subgroup analyses, we will conduct these whenever we consider having a sufficient number of studies providing data for the subgroup analysis.

We will utilise the formal test for subgroup differences [17,37] to evaluate potential differences between subgroups. In case of any significant subgroup difference, we will conduct a separate meta-analysis for each subgroup and present their individual results [17,37]. We have outlined several subgroup analyses below. Given their extensive quantity, there is a risk for both type I and type II errors. Consequently, we will approach the interpretation of our subgroup findings with caution.

- DAA effects in studies at an overall low risk of bias compared to those at moderate, serious, critical, or unknown risk of bias.
- DAA effects in participants without co-morbidities compared to participants with co-morbidities.
- Comparison of individual DAA types to examine any differences.
- The HCV genotypes subgroup compares drug combinations on the same genotype and evaluates each combination’s efficacy on each genotype.
- The IL-28 genotypes subgroup compares drug combinations on the same IL-28 genotype and evaluates each combination’s efficacy on each genotype.
- Comparing geographic regions (Asian compared to non-Asian regions).
- HCV participants according to specific races or ethnicities.
- Study duration. Compare studies by categorising them into two groups based on their follow-up duration: those with a follow-up duration below or equal to the median and those with a follow-up duration above the median. This analysis assesses the influence of shorter follow-up durations in comparison to the median or longer durations.
- Treatment-naive participants compared to previously treated patients.
- DAA effects in participants with chronic kidney disease.
- DAA compared to no DAA in participants coinfected with HIV.
- DAA compared to no DAA in participants coinfected with HBV.
- DAA compared to no DAA in participants with Child-Pugh Class A and B liver cirrhosis.
- DAA compared to no DAA in participants with decompensated liver cirrhosis (on or not on the list for liver transplantation).
- DAA compared to no DAA in liver transplant recipients.
- DAA compared to no DAA in non-hepatic solid organ transplant recipients.
- DAA compared to no DAA in participants with hepatocellular carcinoma.
- DAA compared to no DAA in pregnant women.

### Sensitivity analysis

To address the potential impact of missing data in our analysis, we will conduct the following sensitivity analyses [42].

‘Best-worst-case’ scenario: assuming the best possible and worst possible outcomes for different groups regarding dichotomous outcomes with missing data. For lost-to-follow-up participants in the experimental group, we will assume a scenario where they did not develop a primary outcome event. For participants with missing outcomes in the control group, we will consider scenarios where they developed a primary outcome event.

‘Worst-best-case’ scenario: assuming the best possible and worst possible outcomes for different groups regarding dichotomous outcomes with missing data. For lost-to-follow-up participants in the experimental group, we will assume a scenario where they developed a primary outcome event, and vice versa for the control group.

### Data analysis

We will use a random-effects model, in line with recommendations from previous studies [18,50,53]. This model is appropriate because population characteristics and exposure or outcome definitions often differ across observational studies, and it is rarely justified to assume that all studies estimate the same underlying effect [18]. By accounting for this heterogeneity, a random-effects model can provide a more realistic estimate of the overall effect size.

### Measure of treatment effect

#### Dichotomous outcomes

We plan to conduct meta-analyses to evaluate the efficacy and safety of the interventions under investigation for dichotomous outcomes. If there are zero events reported for a specific outcome, we will utilise the method by Sweeting and colleagues to calculate OR and their corresponding 95% Cis [75]. Additionally, given the anticipated heterogeneity across studies, we will adopt a random-effects model to pool either the RR or OR estimates, depending on the outcome. We will present the results in forest plots.

#### Continuous outcomes

We will conduct a meta-analysis of mean differences for our continuous outcome, which is health-related quality of life scores. For each study, we will extract the mean and standard deviation of the health-related quality of life scores. If the standard deviation is not reported, we will use available data to estimate it, such as the standard error, 95% confidence interval, or interquartile range.

### Trial Sequential Analysis

We will perform Trial Sequential Analysis (TSA)to control the risks of type I and type II errors using the TSA software version 0.9.5.10 Beta developed by the Copenhagen Trial Unit, Centre for Clinical Intervention Research, The Capital Region, Copenhagen University Hospital – Rigshospitalet [78]. TSA has been developed for randomised clinical trials but can also be applied to observational evidence. TSA calculates the required information size and assesses the eventual breach of the cumulative Z-curve of the relevant trial sequential monitoring boundaries for benefit, harm, or futility [10,42,81,82].

We will calculate the required sample size for dichotomous outcomes by considering the proportion of participants in the control group with the outcome, a 20% relative risk reduction, an alpha level of 2.5% or 0.77% for primary and secondary outcomes, respectively; a beta of 10%; and diversity (D^2^) from the meta-analysis[42].In the case of our continuous outcome, we will estimate the required sample size will be estimated based on the standard deviation observed in the control group of studies with low risk of bias, a minimal relevant difference of 50% of this observed standard deviation, an alpha level of 2.5%for primary outcomes; a beta of 10%; and the variance observed in the studies included in the meta-analysis [42].

### GRADE assessments

To evaluate the certainty of evidence and determine the strength of recommendations, we will employ the Grading of Recommendations, Assessment, Development and Evaluation (GRADE) approach. This approach utilises five considerations, namely risk of bias, consistency of effect, imprecision, indirectness, and publication bias, to assess the certainty of evidence for each outcome and to draw conclusions about the certainty of evidence. Our assessments will adhere to the current GRADE guidance, as recommended in Chapter 14 of the Cochrane Handbook for Systematic Reviews of Interventions [72] and in the GRADE Handbook [31,70].

In line with our methodology, we recognise that our application of the GRADE approach will consider the inherent limitations associated with observational studies. We acknowledge that the initial assessment of certainty in the evidence starts at the lowest level (very low), reflecting these inherent limitations [70].

In grading of observational studies, a large intervention effect increases the level of evidence by 1 or 2 levels, and the presence of a dose-response gradient, along with the expectation that all plausible confounding factors would reduce or increase the effect, raises the level of evidence by 1 level.

To evaluate imprecision in our study, we will also utilise TSA. If the accumulated number of participants is less than 33% of the diversity-adjusted required information size (DARIS), we will downgrade the imprecision by three levels in the GRADE analysis. If the number of participants is between 33% - 66% of DARIS, we will downgrade the imprecision by two levels. If the number of participants is between 66% - 100% of DARIS, we will downgrade the imprecision by only one level. However, we will not downgrade if the cumulative Z-curve crosses the monitoring boundaries for benefit, harm, or futility, or if the DARIS is reached. This approach ensures that we consider the precision of our findings in our overall assessment of the certainty of evidence.

### Summary of finding table

We will create ‘Summary of findings’ tables using the GRADEpro Guideline Development Tool (www.gradepro.org). We will display the outcome results on all-cause mortality, serious adverse events, and no SVR because we consider these outcomes critical for decision-makers, i.e. all-cause mortality and serious adverse events outcomes have evident clinical relevance, and SVR having the focus of a potential surrogate outcome in hepatitis C intervention research.We will construct summary of findings table for each class of DAAs used.The protocol for this systematic review and meta-analysis has been registered with PROSPERO under registration number CRD42023494844. It adheres to the Preferred Reporting Items for Systematic Review and Meta-Analysis Protocols [59].

## Discussion

In this protocol, we have outlined our systematic approach to assess the benefits and harms of DAAs in individuals with chronic HCV infection based on observational studies. The study aims to provide a comprehensive analysis of the available observational evidence. We have established inclusion criteria, encompassing various study designs such as quasi-randomised, cohort, case-control, controlled before-and-after, and cross-sectional studies. These criteria ensure a broad scope, allowing us to evaluate the real-world effectiveness and safety of DAA drugs in diverse populations. By including both prospective and retrospective studies, we aim to capture a wide range of data without imposing a minimum duration requirement. Furthermore, we will conduct subgroup analyses to explore potential variations in treatment outcomes based on participant characteristics, comorbidities, DAA types, and other factors, enhancing the generalisability of our findings. The systematic review will also utilise the GRADE approach to assess the quality of evidence, acknowledging the inherent limitations of observational studies. In addition, we will strengthen our analysis using TSA to control the risks of type I and type II errors. This comprehensive approach will provide valuable insights into the role of DAAs in managing chronic HCV infection to inform clinical practice and public health policies.

## Supporting information

Search strategies

## Data Availability

All data produced in the present work are contained in the manuscript

## Abbreviations

ALT: Alanine aminotransferase
AST: Aspartate aminotransferase
CDIM: Clinical diversity in meta-analyses
CI: Confidence interval
DARIS: Diversity-adjusted required information size
DAA: Direct-acting antiviral
GRADE: Grading of Recommendations, Assessment, Development and Evaluation
HBV: Hepatitis B virus
HIV: Human immunodeficiency virus
HCV: Hepatitis C virus
IL-28B: Interleukin 28B
PCR: Polymerase chain reaction
ROBINS-I: Risk of Bias in non-randomized studies of interventions
RR: Relative risk
SVR: Sustained virological response
TSA: Trial Sequential Analysis

## Ethics declarations

Ethics approval and consent to participate

Not applicable

## Consent for publication

Not applicable

## Availability of data and materials

Not applicable

## Acknowledgements

Not applicable

## Authors’ contributions

WMBBK, ATK, GB, DN, RLK, SLK, CG participated in the protocol and study design development. SLK developed the search strategy. WMBBK and CG drafted the manuscript. All authors contributed to the manuscript and approved its final version.

## Footnotes

* As defined by study authors.

