## Supplementary material for "Direct-acting antivirals for chronic hepatitis C infection: a protocol for a systematic review of observational studies": Search strategies

**MEDLINE ALLOvid (1946 to 5 January 2024) (10163 hits)**
1. exp Antiviral Agents/
2. exp Protease Inhibitors/
3. exp Nucleic Acid Synthesis Inhibitors/
4. (direct*acting antiviral* or DAA* or ((protease or polymerase) and inhibitor*) or asunaprevir or beclabuvir or boceprevir or Celuprevir or daklinza or danoprevir or dasabuvir or declatasvir or deleobuvir or elbasvir or exviera or faldaprevir or filibuvir or galexos or grazoprevir or harvoni or incivek or incivo or ledipasvir or mericitabine or narlaprevir or Nesbuvir or odalasvir or olysio or ombitasvir or paritaprevir or radalbuvir or ravidasvir or samatasvir or setrobuvir or simeprevir or sofosbuvir or sovaldi or sovaprevir or sovriad or sunpreva or technivie or tegobuvir or telaprevir or telavic or vaniprevir or vedroprevir or Velpatasvir or victrelis or viekira* or ABT*072 or ABT*450 or ACH*2684 or ACH*2928 or ALS*2200 or BI*201127 or BILB*1941 or BMS*790052 or BMS*986094 or GS*5885 or GS*9256 or GS*0938 or GS*6620 or GS*985* or GSK*2336805 or GSK*2878175 or IDX*184 or IDX*320 or IDX*375 or INN or INX*189 or MK*3281 or MK*3682 or MK*8408 or NPI* or PHX*1766 or PPI*461 or TMC*435 or TMC*647055 or USAN* or VCH*759 or VCH*916 or VX*135 or VX*222 or VX*950).mp.
5. 1 or 2 or 3 or 4
6. exp Hepatitis C, Chronic/
7. (chronic and (hepatitis C or hep C or HCV)).mp.
8. 6 or 7
9. 5 and 8
10. Epidemiologic studies/
11. exp case control studies/
12. exp cohort studies/
13. Case control.tw.
14. (cohort adj (study or studies)).tw.
15. Cohort analy$.tw.
16. (Follow up adj (study or studies)).tw.
17. (observational adj (study or studies)).tw.
18. Longitudinal.tw.
19. Retrospective.tw.
20. Cross sectional.tw.
21. Cross-sectional studies/
22. 10 or 11 or 12 or 13 or 14 or 15 or 16 or 17 or 18 or 19 or 20 or 21
23. 9 and 22

**Embase Ovid (1974 to 5 January 2024)(13750 hits)**
1. exp antivirus agent/
2. exp proteinase inhibitor/
3. exp nucleic acid synthesis inhibitor/
4. (direct*acting antiviral* or DAA* or ((protease or polymerase) and inhibitor*) or asunaprevir or beclabuvir or boceprevir or Celuprevir or daklinza or danoprevir or dasabuvir or declatasvir or deleobuvir or elbasvir or exviera or faldaprevir or filibuvir or galexos or grazoprevir

or harvoni or incivek or incivo or ledipasvir or mericitabine or narlaprevir or Nesbuvir or odalasvir or olysio or ombitasvir or paritaprevir or radalbuvir or ravidasvir or samatasvir or setrobuvir or simeprevir or sofosbuvir or sovaldi or sovaprevir or sovriad or sunpreva or technivie or tegobuvir or telaprevir or telavic or vaniprevir or vedroprevir or Velpatasvir or victrelis or viekira* or ABT*072 or ABT*450 or ACH*2684 or ACH*2928 or ALS*2200 or BI*201127 or BILB*1941 or BMS*790052 or BMS*986094 or GS*5885 or GS*9256 or GS*0938 or GS*6620 or GS*985* or GSK*2336805 or GSK*2878175 or IDX*184 or IDX*320 or IDX*375 or INN or INX*189 or MK*3281 or MK*3682 or MK*8408 or NPI* or PHX*1766 or PPI*461 or TMC*435 or TMC*647055 or USAN* or VCH*759 or VCH*916 or VX*135 or VX*222 or VX*950).mp.

5. 1 or 2 or 3 or 4
6. exp chronic hepatitis C/
7. (chronic and (hepatitis C or hep C or HCV)).mp.
8. 6 or 7
9. 5 and 8
10. Clinical study/
11. Case control study/
12. Family study/
13. Longitudinal study/
14. Retrospective study/
15. Prospective study/
16. Randomized controlled trials/
17. 15 not 16
18. Cohort analysis/
19. (Cohort adj (study or studies)).mp.
20. (Case control adj (study or studies)).tw.
21. (follow up adj (study or studies)).tw.
22. (observational adj (study or studies)).tw.
23. (epidemiologic$ adj (study or studies)).tw.
24. (cross sectional adj (study or studies)).tw.
25. 10 or 11 or 12 or 13 or 14 or 17 or 18 or 19 or 20 or 21 or 22 or 23 or 24
26. 9 and 25

**LILACS (VHL Regional Portal; 1982 to 5 January 2024) (103 hits)**

((mh:(antiviral agents OR d27.505.954.122.388 OR protease inhibitors OR d27.505.519.389.745 OR nucleic acid synthesis inhibitors OR d27.505.519.389.675)) OR (("direct acting antiviral*" OR "direct-acting antiviral" OR daa* OR ((protease OR polymerase) AND inhibitor*) OR asunaprevir OR beclabuvir OR boceprevir OR celuprevir OR daklinza OR danoprevir OR dasabuvir OR declatasvir OR deleobuvir OR elbasvir OR exviera OR faldaprevir OR filibuvir OR galexos OR grazoprevir OR harvoni OR incivek OR incivo OR ledipasvir OR mericitabine OR narlaprevir OR nesbuvir OR odalasvir OR olysio OR ombitasvir OR paritaprevir OR radalbuvir OR ravidasvir OR samatasvir OR setrobuvir OR simeprevir OR sofosbuvir OR sovaldi OR sovaprevir OR sovriad OR sunpreva OR technivie OR tegobuvir OR telaprevir OR telavic OR vaniprevir OR vedroprevir OR velpatasvir OR victrelis OR viekira* OR abt072 OR abt450 OR ach2684 OR ach2928 OR als2200 OR bi201127 OR bilb1941 OR bms790052 OR bms986094 OR gs5885 OR gs9256 OR gs0938 OR gs6620 OR gs985 OR gsk2336805 OR gsk2878175 OR idx184 OR idx320 OR idx375 OR inn OR inx189 OR mk3281 OR mk3682 OR mk8408 OR npi OR phx1766 OR ppi461 OR tmc435 OR tmc647055 OR usan OR vch759 OR vch916 OR vx135 OR vx222 OR vx950 OR abt-072 OR abt-450 OR ach-2684 OR ach-2928 OR als-2200 OR bi-201127 OR bilb-1941 OR bms-790052 OR bms-986094 OR gs-5885 OR gs-9256 OR gs-0938 OR gs-6620 OR gs-985- OR gsk-2336805 OR gsk-2878175 OR idx-184 OR idx-320 OR idx-375 OR inn OR inx-189 OR mk-3281 OR mk-3682 OR mk-8408 OR npi- OR phx-1766 OR ppi-461 OR tmc-435 OR tmc-647055 OR usan- OR vch-759 OR vch-916 OR vx-135 OR vx-222 OR vx-950))) AND ((mh:(hepatitis c, chronic OR c01.221.250.750.120 OR c01.925.440.440.120 OR c01.925.782.350.350.120 OR c06.552.380.350.120 OR c06.552.380.705.440.120 OR c23.550.291.500.477.750)) OR ((chronic AND (hepatitis c OR hep c OR hcv)))) AND ( db:("LILACS"))

**BIOSIS (Web of Science) (1969 to 5 January 2024) (518 hits)**
#5 #4 AND #3
#4 TS=("epidemiologic stud*" or "clinical stud*" or "case control stud*" or "case-control stud*" or "cohort stud*" or "follow up stud*" or "follow-up stud*" or "observational stud*" or "longitudinal stud*" or "retrospective stud*" or "prospective stud*" or "cross sectional stud*" or "cross-sectional stud*" or "family stud*")
#3 #2 AND #1
#2 TS=(chronic and (hepatitis C or hep C or HCV))
#1 TS=(direct*acting antiviral* or DAA* or ((protease or polymerase) and inhibitor*) or asunaprevir OR beclabuvir OR boceprevir OR Celuprevir OR daklinza OR danoprevir OR dasabuvir OR declatasvir OR deleobuvir OR elbasvir OR exviera OR faldaprevir OR filibuvir OR galexos OR grazoprevir OR harvoni OR incivek OR incivo OR ledipasvir OR mericitabine OR narlaprevir OR Nesbuvir OR odalasvir OR olysio OR ombitasvir OR paritaprevir OR radalbuvir OR ravidasvir OR samatasvir OR setrobuvir OR simeprevir OR sofosbuvir OR sovaldi OR sovaprevir OR sovriad OR sunpreva OR technivie OR tegobuvir OR telaprevir OR telavic OR vaniprevir OR vedroprevir OR Velpatasvir OR victrelis OR viekira* OR ABT*072 OR ABT*450 OR ACH*2684 OR ACH*2928 OR ALS*2200 OR BI*201127 OR BILB*1941 OR BMS*790052 OR BMS*986094 OR GS*5885 OR GS*9256 OR GS*0938 OR GS*6620 OR GS*985* OR GSK*2336805 OR GSK*2878175 OR IDX*184 OR IDX*320 OR IDX*375 OR INN OR INX*189 OR MK*3281 OR MK*3682 OR MK*8408 OR NPI* OR PHX*1766 OR PPI*461 OR TMC*435 OR TMC*647055 OR USAN* OR VCH*759 OR VCH*916 OR VX*135 OR VX*222 OR VX*950)

**Science Citation Index Expanded (1900 to 5 January 2024) and Conference Proceedings Citation Index – Science (1990 to 5 January 2024) (Web of Science) (912 hits)**
#5 #4 AND #3
#4 TS=("epidemiologic stud*" or "clinical stud*" or "case control stud*" or "case-control stud*" or "cohort stud*" or "follow up stud*" or "follow-up stud*" or "observational stud*" or "longitudinal stud*" or "retrospective stud*" or "prospective stud*" or "cross sectional stud*" or "cross-sectional stud*" or "family stud*")
#3 #2 AND #1
#2 TS=(chronic and (hepatitis C or hep C or HCV))
#1 TS=(direct*acting antiviral* or DAA* or ((protease or polymerase) and inhibitor*) or asunaprevir OR beclabuvir OR boceprevir OR Celuprevir OR daklinza OR danoprevir OR dasabuvir OR declatasvir OR deleobuvir OR elbasvir OR exviera OR faldaprevir OR filibuvir OR galexos OR grazoprevir OR harvoni OR incivek OR incivo OR ledipasvir OR mericitabine OR narlaprevir OR Nesbuvir OR odalasvir OR olysio OR ombitasvir OR paritaprevir OR radalbuvir OR ravidasvir OR samatasvir OR setrobuvir OR simeprevir OR sofosbuvir OR sovaldi OR sovaprevir OR sovriad OR sunpreva OR technivie OR tegobuvir OR telaprevir OR telavic OR vaniprevir OR vedroprevir OR Velpatasvir OR victrelis OR viekira* OR ABT*072 OR ABT*450 OR ACH*2684 OR ACH*2928 OR ALS*2200 OR BI*201127 OR BILB*1941 OR BMS*790052

OR BMS*986094 OR GS*5885 OR GS*9256 OR GS*0938 OR GS*6620 OR GS*985* OR GSK*2336805 OR GSK*2878175 OR IDX*184 OR IDX*320 OR IDX*375 OR INN OR INX*189 OR MK*3281 OR MK*3682 OR MK*8408 OR NPI* OR PHX*1766 OR PPI*461 OR TMC*435 OR TMC*647055 OR USAN* OR VCH*759 OR VCH*916 OR VX*135 OR VX*222 OR VX*950)
